# Disproportionate increase in cannabis use among individuals with serious psychological distress and association with psychiatric hospitalization and outpatient service use in the National Survey on Drug Use and Health 2009-2019

**DOI:** 10.1101/2023.12.15.23300036

**Authors:** Andrew S Hyatt, Michael William Flores, Benjamin Lê Cooke

## Abstract

**Aims:** Estimate trends in levels of cannabis use among adults with and without serious psychological distress (SPD) in the United States from 2009-2019, and to ascertain whether cannabis use among individuals with SPD was associated with inpatient psychiatric hospitalization and outpatient mental health care.

**Design:** Using multivariable logistic regression models and predictive margin methods, we estimated linear time trends in levels of cannabis use by year and SPD status and rates of psychiatric hospitalization and outpatient service use.

**Setting:** The United States: National Survey on Drug Use and Health (NSDUH), an annual cross-sectional survey, 2009-19 public use files.

**Participants:** 447,228 adults aged ≥ 18 years.

**Measurements:** In the past year, self-report of any and greater-than-weekly cannabis use, any inpatient psychiatric hospitalization, and any outpatient mental health care.

**Findings:** Rates of any and weekly-plus cannabis use increased similarly among individuals with SPD compared to those without from 2009-2014 but more rapidly in SPD from 2015-2019 (p<0.001). Among individuals with SPD, probability of psychiatric hospitalization was greater among individuals with less than weekly (5.2%, 95% CI 4.4-5.9%, p=0.011), and weekly-plus cannabis use (5.4%, 95% CI 4.6-6.1, p=0.002) compared to no use (4.1%, 95% CI 3.8-4.4%). For outpatient mental health care, no use was associated with a 27.4% probability (95% CI 26.7-8.1%) of any outpatient care, significantly less than less than weekly use (32.7% probability, 95% CI 31.3-34.1% p<0.001) and weekly-plus use (29.9% probability, 95% CI 28.3-31.5% p=0.006).

**Conclusions:** Cannabis use is increasing more rapidly among individuals with SPD than the general population, and is associated with increased rates of psychiatric hospitalization as well as increased outpatient service use. These findings can inform policy makers looking to better tailor regulations on advertising for medical and adult use cannabis and develop public health messaging on the use of cannabis in people with mental illness.

## INTRODUCTION

Cannabis is one of the most commonly used substances worldwide, with 147 million individuals using yearly.(1) Rates of cannabis use have been increasing in recent years,(2) a trend that is likely to accelerate given rapidly liberalizing cannabis laws in many jurisdictions. Globally, cannabis use is associated with many fewer harms than legal drugs like alcohol or tobacco,(3) and many individuals can use cannabis without apparent harm.(4) Among populations that are more likely to be harmed by cannabis use are those with mental health disorders.(5) This relationship is most well established with schizophrenia and other psychotic disorders, with cannabis use associated with an increased risk of developing psychosis (6,7) as well as worse outcomes in individuals with the disorder. (8,9) This relationship is less well established with more prevalent mental health conditions, but recent meta-analyses suggest that cannabis use is associated with increased risk of developing anxiety (10) and mood disorders.(11)

While structured diagnostic instruments are not commonly administered in population surveys, shorter measures can provide valid estimates. Serious psychological distress (SPD) has been strongly associated with diagnoses of serious mental illness in the general population (12), poorer overall mental and physical health (13), and greater healthcare utilization compared to those without SPD. (14)

Two recent single year cross sectional studies have found associations between SPD and substance use disorders (15) as well as cannabis use and cannabis use disorder,(16) but given rapid changes in the cannabis regulatory environment longitudinal studies are urgently needed to assess changes in cannabis use over time. One prior study investigated trends over time in cannabis use among individuals with SPD in the United States, and found higher rates of daily use at all time points but no evidence of increasing differences in use between SPD and non-SPD populations from 2008 to 2016. (17)

Additionally, it is not yet known what effects this higher rate of use will have on outcomes among individuals with SPD, nor how cannabis will affect psychiatric service utilization. Psychiatric service use variables provide a useful proxy for mental health and functioning in large community dwelling samples, and there is a large body of literature studying the effect of cannabis on service use in psychosis (18–20). Measurement of outpatient service use can be used to understand whether cannabis is associated with differences in accessing mental health care for patients with high symptom burdens, and inpatient hospitalization is a commonly used proxy for psychiatric relapse and a high overall burden of mental health symptoms. (22,23) There is a very little prior literature investigating the effect of cannabis on hospitalization in other psychiatric disorders (21), and we are unaware of any literature investigating this association in SPD.

The objectives of the present cross-sectional study are twofold. First, we assessed changes over time in the association between SPD and levels of cannabis use, and whether the rate of change in cannabis use was increasing differentially over time. Secondly, we investigated associations between cannabis and outpatient as well as inpatient service use in individuals with SPD in order to understand whether cannabis use was associated with less access to needed outpatient care as well as increased inpatient psychiatric hospitalization, consistent with worse symptoms and functioning. We hypothesized that: 1) there would be a time x SPD interaction such that the rate of cannabis use would increase more rapidly among individuals with SPD compared to those without and 2) that cannabis use would be associated with less outpatient service use as well as greater rates of psychiatric hospitalization among individuals with SPD.

## METHODS

### Participants and study sample

Data were obtained from the 2009-2019 National Survey on Drug Use and Health (NSDUH) public use data files, and was combined across years to allow comparison of cannabis use over time as well as increase precision of service use estimates. The NSDUH is a nationally representative cross-sectional survey that captures respondents’ demographics, substance use, mental health disorders, and health service use. NSDUH collects information from non-institutionalized youth and adults living in the United States and excludes the unhoused, incarcerated, institutionalized, and active-duty military (24). There were a total of 619,411 survey respondents between 2009-2019. The final analytic sample consisted of 444,947, which was all adult respondents ≥ 18 years of age. This study is based on de-identified public use datasets that are exempt from Institutional Review Board review.

### Outcomes

The primary outcome was cannabis use frequency. This was operationalized into two binary indicator variables: any cannabis use in the past year (1 or more days of use) referenced to no use, and weekly-plus cannabis use (>52 days of use in the past year) referenced to less frequent or no use in accordance with past work. (25) The secondary outcomes were outpatient and inpatient mental health service use. Any outpatient mental health service use was operationalized by a respondent indicating they had received any outpatient mental health treatment in the past year, and inpatient psychiatric admission was operationalized by a respondent indicating they had received any inpatient mental health treatment in the past year.

### Independent variables

The main predictor of interest in the primary analysis was past-year serious psychological distress (SPD). This was operationalized using the Kessler Serious Psychological Distress scale (K6), (12) which is a six item instrument assessing how frequently an individual feels hopeless, sad or depressed, nervous, restless or fidgety, like everything is an effort, or feeling down on oneself, no good, worthless. K6 scores ≥13 were assigned a value of 1 indicating the presence of SPD in the past year, and score <13 were assigned a value of 0 indicating no past year SPD. This cutoff has been shown to be an optimal indicator for the presence of clinically significant psychiatric disorder.(12,26) Given interest in changes in substance use over time, year of survey response was also entered into the model.

In the secondary analysis of service use in individuals with any past year SPD, cannabis use was used as a predictor in the analysis of psychiatric service use. This was operationalized into a single three level variable, with levels of no use, less than weekly use (1-52 days of use in the past year), and weekly-plus use (>52 days of use in the past year).

Regression models adjusted for covariates known to confound the relationship between SPD, cannabis use, and psychiatric service use.(14,17,27,28) Covariates included race and ethnicity (white, non-Hispanic Black, Hispanic, non-Hispanic other), sex (male, female), yearly income (<$20,000, $20,000-49,999, $50,000-74,999, $75,000 or more), age (18-25 or 26 and older), marital status (currently married, widowed, separated or divorced, never married), education (less than high school, high school graduate, some college, or college graduate), and heavy alcohol use (yes/no). Models estimating psychiatric service use also adjusted for health insurance (private health insurance, Medicaid, Medicare, other insurance, or uninsured).

After the above models were analyzed, we conducted a post-hoc exploratory analysis assessing whether changes in risk perceptions mediated differences in cannabis use by SPD status. For any cannabis use, participants were asked what the risk of monthly cannabis use was, and for weekly plus cannabis use, participants were asked what the risk of using cannabis one or two times per week. Both measures were dichotomized into “perceived great risk” and “other perceived risk” in accordance with past research (29,30) and entered into the models as above.

### Statistical analysis

We used chi-squared tests to compare baseline differences in the outcomes and predictors between adults with and without SPD. Unadjusted rates of cannabis use by SPD over time were plotted. Next, we estimated two multivariable logistic regression models with an interaction term between year and SPD, first on any cannabis use and then on weekly-plus cannabis use, conditional on the above covariates, to allow estimation of changes in the relationship between SPD and cannabis use over time in the full dataset (*n*=444,947).

Next, to assess the impact of level of cannabis use on psychiatric service utilization, we estimated multivariable logistic regression models to predict any inpatient psychiatric hospitalization and any outpatient service use among the sub-population of individuals with past-year serious psychological distress (*n*=64,137).

To interpret interaction terms in all non-linear models, predictive margins methods were used decompose interaction terms as well as allow for conversion of regression coefficients to predicted probabilities for interpretability.(31)

Sampling weights were computed to control for non-response and were adjusted for consistency with other data from the U.S. Census Bureau. In accordance with instructions published in the NSDUH public use data files, after aggregating the 11 data sets a new weight was calculated by dividing the original weight by the number of data sets involved. (32) Results were considered statistically significant at p<0.05 (two tailed). All analyses were conducted in Stata, version 16.(33)

## RESULTS

### Descriptive results by SPD

Individuals with past year SPD had higher rates of any cannabis use (27.4% vs 11.7%) and weekly-plus cannabis use (13.9% vs 5.6%) compared to individuals without past year SPD. There were significant differences between groups in all demographic (e.g., sex, race, ethnicity) and socioeconomic (e.g. income, insurance) characteristics (Table 1).

**Table 1:** Demographic characteristics and cannabis use by past year serious psychological distress (SPD)

|  | No past year SPD | Past year SPD |  |
| --- | --- | --- | --- |
| **Cannabis use** |  |  | p<0.001 |
| none | 88.34% | 72.58% |  |
| any cannabis use | 11.66% | 27.42% |  |
| weekly-plus cannabis | 5.56% | 13.86% |  |
| **Past year psychiatric care received** |  |  |  |
| 1+ inpatient hospitalization | 0.45% | 4.49% | p<0.001 |
| 1+ outpatient mental health visit | 4.42% | 28.60% |  |
| **Heavy alcohol use** |  |  | p<0.001 |
| No heavy alcohol use | 93.54% | 90.27% |  |
| Heavy alcohol use | 6.46% | 9.73% |  |
| **Race** |  |  | p<0.001 |
| White, non-Hispanic | 65.51% | 67.64% |  |
| Black, non-Hispanic | 11.88% | 11.17% |  |
| Hispanic | 15.41% | 14.75% |  |
| non-Hispanic, other | 7.20% | 6.44% |  |
| **Sex** |  |  | p<0.001 |
| Male | 49.46% | 38.17% |  |
| Female | 50.54% | 61.83% |  |
| **Total family income** |  |  | p<0.001 |
| Less than \$20,000 | 16.23% | 27.70% | |
| \$20,000 - \$49,999 | 30.76% | 32.90% | |
| \$50,000 - \$74,999 | 16.70% | 14.73% | |
| \$75,000 or More | 36.31% | 24.67% | |
| **Age** |  |  | p<0.001 |
| 18-25 | 12.76% | 27.20% |  |
| 26 or older | 87.24% | 72.80% |  |
| **Insurance coverage** |  |  | p<0.001 |
| Private health insurance | 54.51% | 48.94% |  |
| Medicaid | 8.39% | 17.69% |  |
| Medicare | 22.46% | 13.52% |  |
| Other health insurance | 2.00% | 2.62% |  |
| Uninsured | 12.64% | 17.23% |  |
| **Marital status** |  |  | p<0.001 |
| Married | 54.64% | 33.05% |  |
| Widowed | 6.18% | 4.13% |  |
| Divorced or Separated | 13.48% | 18.03% |  |
| Never Been Married | 25.70% | 44.78% |  |
| **Education level** |  |  | p<0.001 |
| Less than high school | 13.30% | 15.38% |  |
| High school graduate | 27.45% | 28.23% |  |
| Some college | 27.84% | 33.15% |  |
| College graduate | 31.41% | 23.25% |  |

### Trends in past year cannabis use by SPD

#### Any past year cannabis use

Unadjusted rates of past year cannabis use can be found in Figure 1. After adjusting for covariates, rates of cannabis use among individuals with and without SPD rose similarly from 2009-2014, but use in SPD rose significantly faster in 2015 (1.70% increase in SPD, 95% CI 0.006-3.4%, p=0.049), and remained significant through 2019 (5.0% increase in SPD, 95% CI 2.8-7.2%, p<0.001), indicating that rates of any cannabis use increased faster among individuals with SPD compared to those without during this time period indexed to 2009.

**Figure 1.**
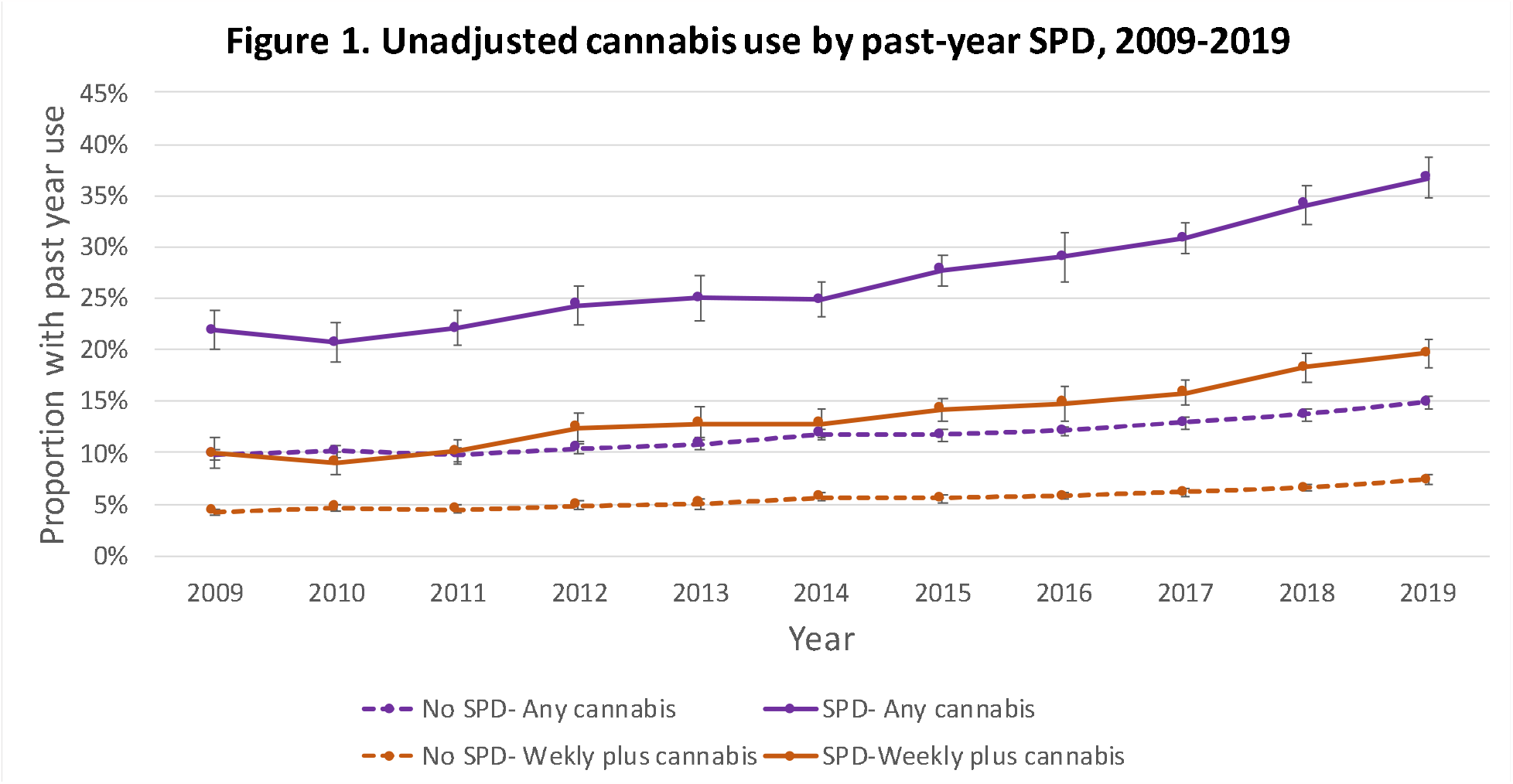
Unadjusted prevalence of past year of any and weekly-plus cannabis use, stratified by past year serious psychological distress (SPD)

#### Weekly-plus past year cannabis use

After adjusting for covariates, rates of cannabis use among individuals with and without SPD rose similarly from 2009-2014, but use in SPD rose again significantly faster in 2015 (1.6% additional increase in SPD group, 95%. CI 0.3-2.9%, p=0.014) and remained significant through 2019 (3.5% additional increase in SPD group, 95% CI 2.0-5.0%, p<0.001), indicating that rates of weekly-plus cannabis use increased faster among individuals with SPD compared to those without during this time period indexed to 2009. See online appendix for full regression results and comparisons of predicted probabilities

### Effect of cannabis use on psychiatric service utilization in SPD

#### Past year inpatient psychiatric hospitalization

After adjusting for covariates, among individuals with SPD no past year cannabis use was associated with a 4.1% probability of hospitalization (95% CI 3.8-4.4%), less than weekly use with a 5.2% probability (95% CI 4.4-5.9%), and weekly-plus use with a 5.4% probability (95% CI 4.6-6.1%, see Figure 2). Compared to no use, risk of hospitalization was significantly higher in both less than weekly use (p=0.011) and weekly-plus use (p=0.002). There was no significant difference in hospitalization risk between less than weekly use and weekly plus use (p=0.67)

**Figure 2.**
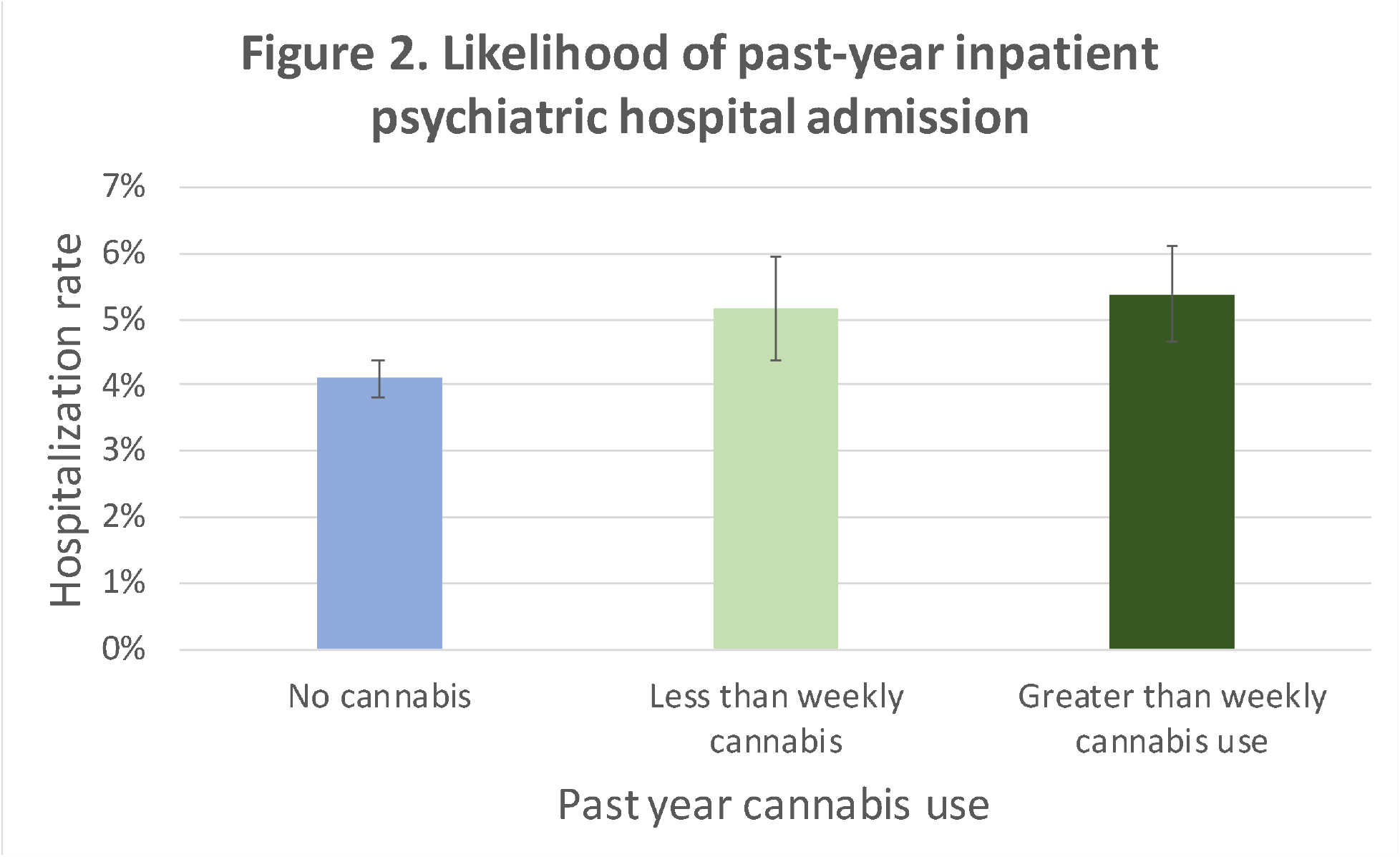
Adjusted likelihood of any past-year inpatient psychiatric hospitalization 2009-2019 among individuals with serious psychological distress (SPD), stratified by cannabis use

#### Past year outpatient psychiatric care

After adjusting for covariates, no cannabis use was associated with a 27.3% probability of any outpatient mental health care (95% CI 26.7-28.1%), less than weekly use was associated with a 32.6% probability (95% CI 31.3-34.1%) and weekly-plus use was associated with a 29.9% probability (95% CI 28.3-31.5%, see figure 3). Both less than weekly (p=0.002) and weekly-plus users (p=0.006) had a higher probability of receiving any outpatient care than non-users, and less than weekly users were significantly more likely to have an outpatient visit than less-than weekly-plus users (p=0.013). See online appendix for full regression results.

**Figure 3.**
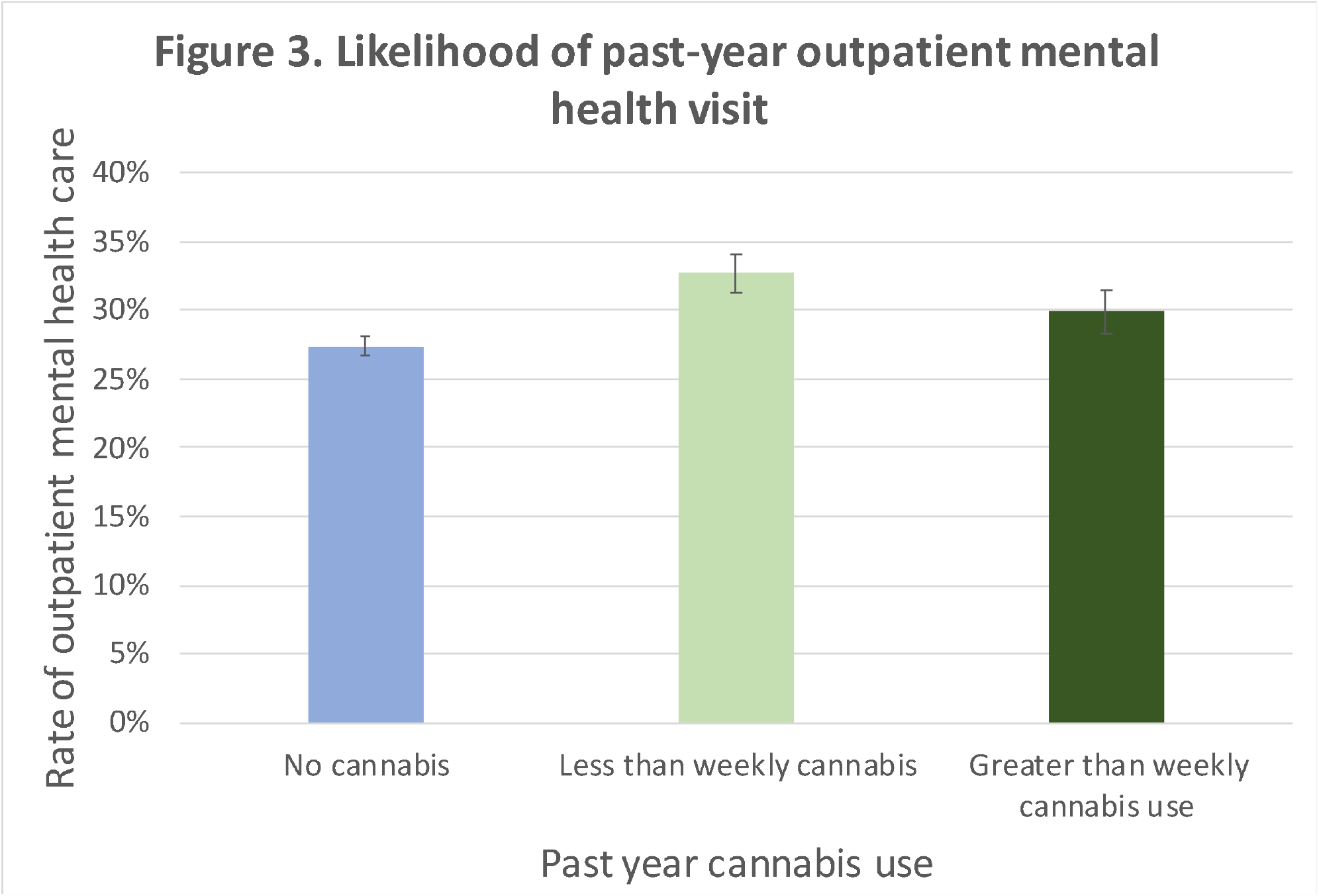
Adjusted likelihood of any past-year outpatient mental health service use 2009-2019 among individuals with serious psychological distress (SPD), stratified by cannabis use

### Exploratory analysis of the effect of risk perceptions on differences in cannabis use

To ascertain whether risk perceptions could partially explain changes in cannabis use by SPD status, both regression models investigating cannabis use over time by SPD were run with the additional of perceived risk of monthly cannabis use (for use of any cannabis) and use 1-2 times per week (weekly plus cannabis). Inclusion of risk perceptions attenuated but did not eliminate the relationship between SPD and cannabis use such that in both models the difference in changes in rates use between SPD and no SPD was only significant in 2018 and 2019 (results available upon request).

## DISCUSSION

The present study found that cannabis use has been accelerating more quickly in individuals with SPD compared to those without since 2015, with similar changes in any use and greater than weekly use. Furthermore, cannabis use among individuals with SPD was associated with higher rates of inpatient hospitalization as well as increases in outpatient psychiatric service use. This suggests that cannabis use may be associated with a higher burden of psychiatric illness given higher rates of hospitalization as well as more help seeking behavior as indicated by higher levels of outpatient service use. This should be considered by policy makers implementing regulations on medical cannabis, which is often available for mental health concerns, as well as for public health messaging and advertising regulations for recreational cannabis.

We found that both any use and weekly-plus cannabis use increased more rapidly among individuals with SPD in the second half of the study period (2015-2019), which contrasts with one prior study that found no interaction between SPD and year in cannabis use up to 2016. (17) While, this prior study used different categories to break down cannabis use making it difficult to compare results directly, the consistent association across levels of cannabis use in the present study and more recent follow-up make it likely that this reflects true alterations in patterns of use among individuals with SPD in recent years. A similar study among adults with depression found a more rapid increase in cannabis use among those with depression compared to those without from 2005-2017, a finding that was associated with a larger drop in risk perception among individuals with depression. (29)

Lower risk perceptions have been tied to higher rates cannabis use in the general population (34), and a recent analysis of a similar time period to the present study found that following recreational cannabis legalization use among young adults increased and the association of lower perception of harms with higher rates of use was strengthened. (35) Supporting this interpretation, the inclusion of risk perception in an exploratory analysis reduced the association between SPD and more rapid increases in cannabis use, although it did not eliminate it. This suggests that changing risk perceptions may partially explain the differences in use between those with SPD and those without. Marketing touting the relative harmlessness of cannabis as well as its utility in treating mental illness is common (36) but poorly supported by research.(37) In the present study, trends began to diverge in 2015, when only two states had legalized recreational cannabis, but 23 had legalized cannabis for medical use.(38) Prior research has shown higher rates of cannabis use in medical cannabis states and among individuals with SPD but no interaction between medical cannabis laws and SPD,(39) suggesting that the presence of medical cannabis laws alone do not fully explain this relationship. A recent study of recreational cannabis marketing in California found that high density of marketing promoting the health benefits of cannabis was associated with a 2-3x increased odds of cannabis use among adults exposed to health messaging.(40) Given the rapidly increasing number of states with legal recreational cannabis, future research should investigate the effect of cannabis commercialization, availability, and health-related marketing on use among individuals with SPD and other serious mental illness.

The present findings also indicate that among individuals with SPD, cannabis use at any level is associated with higher rates of inpatient hospitalization, consistent with more severe mental health symptoms. Previous research has shown that cannabis use disorder has been associated with more emergency department visits and medical inpatient hospitalizations, (28,41) but there has been less focus on associations with psychiatric hospitalization. One Dutch study found a dose-dependent association between cannabis use and inpatient psychiatric admission, (21) and a single year NSDUH analysis from 2007 found an increased risk of psychiatric hospitalization among individuals using both alcohol and cannabis, but not among people only using cannabis.(42) Of note the Dutch study did not control for alcohol use as the present analysis did, which is critical given that much prior research on the relationship between cannabis and mental health outcomes has been criticized for not controlling for alcohol use.(43) Given the robust set of covariates used to control for confounding in the present analysis as well as the multi-year dataset analyzed, the present study provides stronger support for the relationship between cannabis use and psychiatric hospitalization among individuals with serious mental illness.

There was also an association between cannabis use and outpatient service utilization, with weekly-plus users utilizing more outpatient psychiatric care than non-users, and less than weekly users utilizing significantly more care than both groups. This result ran contrary to our hypothesis that cannabis use would be associated with less engagement with outpatient care as seen in the psychosis literature, (20,44) and suggests that cannabis use in the SPD population could be associated with more help seeking behavior as demonstrated by the increased likelihood of any outpatient mental health care. Given past research suggesting falling rates of substance use disorder treatment among individuals using cannabis, especially in states that have legalized cannabis,(35) these results suggest that general outpatient mental health clinics will become increasingly important in treating individuals with comorbid mental illness and cannabis use.

The present study has several limitations. First, given all responses were by self-report, respondents’ answers could not be verified, and could be subject to social desirability bias. Use of computer assisted interviews may reduce this in the NSDUH. Secondly, given the NSDUH’s cross-sectional nature we could not infer the direction of the causal relationship between cannabis and increased levels of inpatient and outpatient service use. Future research using longitudinal data could more easily ascertain this effect on individuals, and location specific data not available in the public-use NSDUH files could be used to assess the impact of recreational and medical cannabis laws on both cannabis use and psychiatric services. Finally, the potency of cannabis has increased markedly over time as has availability of different routes of administration, and there is good evidence that high potency use may be associated with worse outcomes in people with mental illness. (45–47) We were unable to ascertain this relationship in the NSDUH as this dataset does not collect information on potency or route of administration, and future research should investigate this important mediator of the relationship between cannabis use and mental health outcomes as a possible way of mitigating harms of cannabis legalization.

## CONCLUSIONS

Our findings demonstrate that use of cannabis is increasing more rapidly among individuals with SPD vs those without, suggesting that individuals with probable serious mental illness may be more sensitive to changing cultural norms around cannabis compared to the general population. Additionally, among individuals with SPD, cannabis use was associated with more inpatient hospitalizations, suggesting that this increased use is also correlated with a higher burden of mental health symptoms. Outpatient service use was also higher in individuals with SPD and cannabis use, which indicates that general outpatient clinics can be an effective site of intervention for individuals with comorbid cannabis use and mental illness. Future research should investigate the effect of changing cannabis regulations, especially cannabis marketing and health claims, to inform cannabis regulations to optimize public health.

## Data Availability

All data are publicly available through the National Survey of Drug Use and Health public use files.

https://www.datafiles.samhsa.gov/dataset/national-survey-drug-use-and-health-2019-nsduh-2019-ds0001

## Notes

### Competing Interest Statement

The authors have declared no competing interest.

### Funding Statement

The work was supported by the Dupont-Warren Fellowship from the Harvard Medical School Department of Psychiatry as well as the Cambridge Health Alliance Population Health Scholars Program

