## Supplemental Tables for "Disproportionate increase in cannabis use among individuals with serious psychological distress and association with psychiatric hospitalization and outpatient service use in the National Survey on Drug Use and Health 2009-2019"

**Model 1: Full regression results for any cannabis use by SPD and year**

**Survey: Logistic regression**

|  | OR | p-value | [95% Conf | Interval] |
| --- | --- | --- | --- | --- |
| Year (ref=2009) |  |  |  |  |
| 2010 | 1.039 | .364 | .951 | 1.134 |
| 2011 | 1.014 | .7 | .939 | 1.096 |
| 2012 | 1.085 | .063 | .995 | 1.183 |
| 2013 | 1.145 | .002 | 1.066 | 1.23 |
| 2014 | 1.287 | <0.001 | 1.198 | 1.382 |
| 2015 | 1.289 | <0.001 | 1.192 | 1.394 |
| 2016 | 1.336 | <0.001 | 1.247 | 1.433 |
| 2017 | 1.48 | <0.001 | 1.375 | 1.593 |
| 2018 | 1.602 | <0.001 | 1.482 | 1.732 |
| 2019 | 1.831 | <0.001 | 1.699 | 1.972 |
| Any past year SPD | 2.105 | <0.001 | 1.853 | 2.392 |
| Year x SPD interaction (ref=2009) | 1 | . | . | . |
| 2010 | .88 | .184 | .722 | 1.073 |
| 2011 | 1.003 | .978 | .82 | 1.226 |
| 2012 | 1.074 | .444 | .88 | 1.311 |
| 2013 | 1.065 | .431 | .899 | 1.262 |
| 2014 | .897 | .195 | .754 | 1.067 |
| 2015 | 1.039 | .595 | .892 | 1.21 |
| 2016 | 1.069 | .435 | .892 | 1.282 |
| 2017 | 1.005 | .942 | .861 | 1.173 |
| 2018 | 1.141 | .115 | .963 | 1.351 |
| 2019 | 1.136 | .136 | .954 | 1.352 |
| Heavy alcohol use | 3.974 | <0.001 | 3.823 | 4.132 |
| Race (reference=White) |  |  |  |  |
| 2 – Non-Hispanic Black | .937 | .008 | .897 | .98 |
| 3 – Non-Hispanic Native American | 1.247 | .003 | 1.095 | 1.42 |
| 4 – Non-Hispanic Native Pacific Islander | .819 | .076 | .653 | 1.026 |
| 5 – Non-Hispanic Asian | .381 | <0.001 | .353 | .411 |
| 6 – Non-Hispanic other | 1.457 | <0.001 | 1.323 | 1.605 |
| 7 - Hispanic | .662 | <0.001 | .633 | .692 |
| Female sex | .626 | <0.001 | .609 | .644 |
| Income (ref=$75,000 or more) |  |  |  |  |
| Less than $20,000 | 1.141 | <0.001 | 1.096 | 1.188 |
| $20,000 - $49,999 | 1.008 | .675 | .969 | 1.048 |
| $50,000 - $74,999 | .945 | .022 | .903 | .99 |
| Age >25 | .528 | <0.001 | .511 | .546 |
| Marital status (ref=married) |  |  |  |  |
| Widowed | .583 | <0.001 | .522 | .651 |
| Divorced or Separated | 1.783 | <0.001 | 1.696 | 1.874 |
| Never Been Married | 3.266 | <0.001 | 3.135 | 3.401 |
| Education (ref=college graduate) |  |  |  |  |
| Less than high sch~l | .886 | .001 | .835 | .941 |
| High school graduate | .896 | <0.001 | .855 | .938 |
| Some college | 1.096 | .001 | 1.051 | 1.144 |

Predicted probability of any cannabis use by year x SPD

Expression	: Pr(any_cannabis), predict()

-----------------------------------------------------------------------------------------

| Delta-method

| Margin Std. Err. t P>|t| [95% Conf. Interval]

------------------------+----------------------------------------------------------------

year#spdyr |

2009#No past year SPD |	.1001507	.0019797	50.59	0.000	.0957934	.1045081

2009#Any past year SPD |	.1759327	.0066879	26.31	0.000	.1612128	.1906525

2010#No past year SPD |	.103222	.0026518	38.93	0.000	.0973855	.1090585

2010#Any past year SPD |	.1649675	.0072492	22.76	0.000	.149012	.180923

2011#No past year SPD |	.1012718	.0027714	36.54	0.000	.0951721	.1073716

2011#Any past year SPD |	.1780058	.0061113	29.13	0.000	.164555	.1914566

2012#No past year SPD |	.1068424	.0023923	44.66	0.000	.101577	.1121078

2012#Any past year SPD |	.1957954	.0071363	27.44	0.000	.1800884	.2115023

2013#No past year SPD |	.1115049	.0022664	49.20	0.000	.1065165	.1164933

2013#Any past year SPD |	.2019833	.0075015	26.93	0.000	.1854726	.218494

2014#No past year SPD |	.1221433	.0019697	62.01	0.000	.117808	.1264787

2014#Any past year SPD |	.1944985	.0061145	31.81	0.000	.1810406	.2079563

2015#No past year SPD |	.1222737	.002395	51.05	0.000	.1170023	.1275452

2015#Any past year SPD |	.2151336	.0054747	39.30	0.000	.2030838	.2271833

2016#No past year SPD |	.1257361	.0018751	67.06	0.000	.121609	.1298631

2016#Any past year SPD |	.2246617	.0087824	25.58	0.000	.2053317	.2439917

2017#No past year SPD |	.1358768	.002267	59.94	0.000	.1308872	.1408664

2017#Any past year SPD |	.2306694	.0054297	42.48	0.000	.2187187	.2426202

2018#No past year SPD |	.1442374	.0026661	54.10	0.000	.1383693	.1501054

2018#Any past year SPD |	.2631134	.0072014	36.54	0.000	.2472631	.2789637

2019#No past year SPD |	.1591159	.0025989	61.22	0.000	.1533957	.1648362

2019#Any past year SPD |	.2847859	.0082839	34.38	0.000	.266553	.3030187

-----------------------------------------------------------------------------------------

Pairwise comparisons of average marginal effects for model 1
Expression	: Pr(any_cannabis), predict()
dy/dx w.r.t. : 2010.year 2011.year 2012.year 2013.year 2014.year 2015.year 2016.year 2017.year 2018.year 2019.year

|  | Adjusted difference | Std.Err. | | p | [95%Conf. | Interval] |
| --- | --- | --- | --- | --- | --- | --- |
| 2009 (reference) | | |  |  |  |  |
| 2010 |  |  | |  |  |  |
| Any past year SPD vs No past year SPD | -0.014 | 0.010 | | 0.173 | -0.034 | 0.006 |
| 2011 | | |  |  |  |  |
| Any past year SPD vs No past year SPD | 0.001 | 0.011 | | 0.930 | -0.020 | 0.022 |
| 2012 | | |  |  |  |  |
| Any past year SPD vs No past year SPD | 0.013 | 0.011 | | 0.229 | -0.008 | 0.035 |
| 2013 | | |  |  |  |  |
| Any past year SPD vs No past year SPD | 0.015 | 0.010 | | 0.135 | -0.005 | 0.034 |
| 2014 | | |  |  |  |  |
| Any past year SPD vs No past year SPD | -0.003 | 0.010 | | 0.723 | -0.023 | 0.016 |
| 2015 | | |  |  |  |  |
| Any past year SPD vs No past year SPD | 0.017 | 0.009 | | 0.049 | 0.000 | 0.034 |
| 2016.year | | |  |  |  |  |
| spdyr | | |  |  |  |  |
| Any past year SPD vs No past year SPD | 0.023 | 0.011 | | 0.037 | 0.001 | 0.045 |
| 2017.year | | |  |  |  |  |
| spdyr | | |  |  |  |  |
| Any past year SPD vs No past year SPD | 0.019 | 0.009 | | 0.033 | 0.002 | 0.036 |
| 2018.year | | |  |  |  |  |
| spdyr | | |  |  |  |  |
| Any past year SPD vs No past year SPD | 0.043 | 0.010 | | 0.000 | 0.023 | 0.064 |
| 2019.year | | |  |  |  |  |
| spdyr | | |  |  |  |  |
| Any past year SPD vs No past year SPD | 0.050 | 0.011 | | 0.000 | 0.028 | 0.072 |

**Model 2: Full regression results for weekly-plus cannabis use by SPD and year**

**Survey: Logistic regression**

|  | OR | p-value | [95% Conf | Interval] |
| --- | --- | --- | --- | --- |
| Year (ref=2009) |  |  |  |  |
| 2010 | 1.102 | .11 | .974 | 1.247 |
| 2011 | 1.089 | .128 | .972 | 1.22 |
| 2012 | 1.175 | .026 | 1.024 | 1.349 |
| 2013 | 1.226 | .003 | 1.089 | 1.381 |
| 2014 | 1.437 | <0.001 | 1.297 | 1.592 |
| 2015 | 1.414 | <0.001 | 1.262 | 1.585 |
| 2016 | 1.492 | <0.001 | 1.349 | 1.651 |
| 2017 | 1.642 | <0.001 | 1.474 | 1.828 |
| 2018 | 1.775 | <0.001 | 1.61 | 1.957 |
| 2019 | 2.06 | <0.001 | 1.85 | 2.294 |
| Any past year SPD | 1.926 | <0.001 | 1.625 | 2.284 |
| Year x SPD interaction (ref=2009) |  |  |  |  |
| 2010 | .802 | .077 | .624 | 1.029 |
| 2011 | .941 | .631 | .717 | 1.234 |
| 2012 | 1.112 | .386 | .859 | 1.439 |
| 2013 | 1.132 | .313 | .874 | 1.465 |
| 2014 | .939 | .54 | .752 | 1.171 |
| 2015 | 1.083 | .439 | .87 | 1.348 |
| 2016 | 1.061 | .593 | .836 | 1.347 |
| 2017 | 1.032 | .75 | .834 | 1.277 |
| 2018 | 1.188 | .083 | .973 | 1.449 |
| 2019 | 1.129 | .237 | .912 | 1.398 |
| Heavy alcohol use | 3.246 | <0.001 | 3.088 | 3.411 |
| Race (reference=White) |  |  |  |  |
| 2 – Non-Hispanic Black | 1.103 | .001 | 1.048 | 1.162 |
| 3 – Non-Hispanic Native American | 1.217 | .012 | 1.054 | 1.405 |
| 4 – Non-Hispanic Native Pacific Islander | .78 | .111 | .569 | 1.069 |
| 5 – Non-Hispanic Asian | .313 | <0.001 | .27 | .363 |
| 6 – Non-Hispanic other | 1.582 | <0.001 | 1.424 | 1.758 |
| 7 - Hispanic | .629 | <0.001 | .595 | .664 |
| Female sex | .504 | <0.001 | .485 | .524 |
| Income (ref=$875000 or more) |  |  |  |  |
| Less than $20,000 | 1.339 | <0.001 | 1.26 | 1.422 |
| $20,000 - $49,999 | 1.247 | <0.001 | 1.184 | 1.313 |
| $50,000 - $74,999 | 1.123 | .003 | 1.052 | 1.199 |
| Age >25 | .634 | <0.001 | .611 | .659 |
| Marital status (ref=married) |  |  |  |  |
| Widowed | .608 | <0.001 | .521 | .71 |
| Divorced or Separated | 1.764 | <0.001 | 1.643 | 1.894 |
| Never Been Married | 3.121 | <0.001 | 2.945 | 3.308 |
| Education (ref=college graduate) |  |  |  |  |
| Less than high sch~l | 1.58 | <0.001 | 1.465 | 1.705 |
| High school graduate | 1.53 | <0.001 | 1.436 | 1.63 |
| Some college | 1.59 | <0.001 | 1.494 | 1.692 |

Adjusted probability of weekly-plus cannabis use

Expression	: Pr(weeklyplus_cannabis), predict()

-----------------------------------------------------------------------------------------

| Delta-method

| Margin Std. Err. t p [95% Conf. Interval]

------------------------+----------------------------------------------------------------

year#spdyr |

2009#No past year SPD |	.0429694	.0013475	31.89	0.000	.0400036	.0459352

2009#Any past year SPD |	.0993362	.0066328	14.98	0.000	.0847375	.113935

2010#No past year SPD |	.0468928	.0015884	29.52	0.000	.0433967	.0503889

2010#Any past year SPD |	.0897318	.0053135	16.89	0.000	.0780369	.1014267

2011#No past year SPD |	.0452898	.0016077	28.17	0.000	.0417513	.0488283

2011#Any past year SPD |	.1005996	.0058082	17.32	0.000	.0878159	.1133834

2012#No past year SPD |	.0491857	.0018956	25.95	0.000	.0450134	.053358

2012#Any past year SPD |	.1240315	.0067255	18.44	0.000	.1092289	.1388342

2013#No past year SPD |	.05066	.0018908	26.79	0.000	.0464984	.0548215

2013#Any past year SPD |	.1281691	.0073058	17.54	0.000	.1120892	.1442491

2014#No past year SPD |	.0574901	.0014506	39.63	0.000	.0542974	.0606829

2014#Any past year SPD |	.128028	.0062275	20.56	0.000	.1143214	.1417346

2015#No past year SPD |	.0558139	.0018528	30.12	0.000	.0517359	.0598918

2015#Any past year SPD |	.1416883	.0052138	27.18	0.000	.1302128	.1531638

2016#No past year SPD |	.0582163	.0012745	45.68	0.000	.0554111	.0610214

2016#Any past year SPD |	.1479811	.0078632	18.82	0.000	.1306743	.1652879

2017#No past year SPD |	.0623311	.0018808	33.14	0.000	.0581916	.0664707

2017#Any past year SPD |	.1578123	.005576	28.30	0.000	.1455396	.170085

2018#No past year SPD |	.0663622	.0015042	44.12	0.000	.0630514	.069673

2018#Any past year SPD |	.1826497	.0064017	28.53	0.000	.1685596	.1967398

2019#No past year SPD |	.073815	.0021131	34.93	0.000	.0691642	.0784659

2019#Any past year SPD |	.1967222	.0061477	32.00	0.000	.1831912	.2102533

-----------------------------------------------------------------------------------------

Pairwise comparisons of average marginal effects

|  | Adjusted difference | Std.Err. | | | P>t | [95%Conf. | | Interval] |
| --- | --- | --- | --- | --- | --- | --- | --- | --- |
| 2009 (base) | | |  | | | |  |  |
| 2010 | | | |  |  |  |  |  |
| Any past year SPD vs No past year SPD | -0.012 | 0.006 | | | 0.077 | -0.024 | | 0.001 |
| 2011 | | | |  |  |  |  |  |
| Any past year SPD vs No past year SPD | -0.002 | 0.007 | | | 0.805 | -0.017 | | 0.013 |
| 2012 | | | |  |  |  |  |  |
| Any past year SPD vs No past year SPD | 0.012 | 0.008 | | | 0.117 | -0.003 | | 0.028 |
| 2013 | | | |  |  |  |  |  |
| Any past year SPD vs No past year SPD | 0.015 | 0.008 | | | 0.068 | -0.001 | | 0.031 |
| 2014 | | | |  |  |  |  |  |
| Any past year SPD vs No past year SPD | 0.005 | 0.007 | | | 0.453 | -0.008 | | 0.018 |
| 2015 | | | |  |  |  |  |  |
| Any past year SPD vs No past year SPD | 0.016 | 0.007 | | | 0.014 | 0.003 | | 0.029 |
| 2016 | | | |  |  |  |  |  |
| Any past year SPD vs No past year SPD | 0.017 | 0.008 | | | 0.038 | 0.001 | | 0.032 |
| 2017 | | | |  |  |  |  |  |
| Any past year SPD vs No past year SPD | 0.017 | 0.007 | | | 0.011 | 0.004 | | 0.031 |
| 2018 | | | |  |  |  |  |  |
| Any past year SPD vs No past year SPD | 0.034 | 0.006 | | | 0.000 | 0.021 | | 0.047 |
| 2019 | | | |  |  |  |  |  |
| Any past year SPD vs No past year SPD | 0.035 | 0.007 | | | 0.000 | 0.020 | | 0.050 |
